# Converting dose-area product to effective dose in dental cone-beam computed tomography using organ-specific deep learning

**DOI:** 10.1101/2024.05.28.24308014

**Authors:** Ruben Pauwels

## Abstract

**Objective:** To develop an accurate method for converting dose-area product (DAP) to patient dose for dental cone-beam computed tomography (CBCT) using deep learning.

**Methods:** 24,384 CBCT exposures of an adult phantom were simulated with PCXMC 2.0, using permutations of tube voltage, filtration, source-isocenter distance, beam width/height and isocenter position. Equivalent organ doses as well as DAP values were recorded. Next, using the aforementioned scan parameters as inputs, neural networks (NN) were trained using Keras for estimating the equivalent dose per DAP for each organ. Two methods were explored for positional input features: (1) ‘Coordinate’ mode, which uses the (continuous) XYZ-coordinates of the isocenter, and (2) ‘AP/JAW’ mode, which uses the (categorical) anteroposterior and craniocaudal position. Each network was trained, validated and tested using a 3/1/1 data split. Effective dose (ED) was calculated from the combination of NN outputs using ICRP 103 tissue weighting factors. The performance of the resulting NN models for estimating ED/DAP was compared with that of a multiple linear regression (MLR) model as well as direct conversion coefficients (CC).

**Results:** The mean absolute error (MAE) for organ dose / DAP on the test data ranged from 0.18% (bone surface) to 2.90% (oesophagus) in ‘Coordinate’ mode and from 2.74% (red bone-marrow) to 14.13% (brain) in ‘AP/JAW’ mode. The MAE for ED was 0.23% and 4.30%, respectively, for the two modes, vs. 5.70% for the MLR model and 20.19%-32.67% for the CCs.

**Conclusion:** NNs allow for an accurate estimation of patient dose based on DAP in dental CBCT.

## Introduction

Since the introduction of cone-beam computed tomography (CBCT) into dentistry over 25 years ago, its patient dose has been a continuous topic of interest.^1^ Whereas effective doses in dentistry tend to be relatively low due to the limited amount of radiosensitive tissue in the head and neck,^2,3^ dental imaging is among the more frequent types of medical imaging, resulting in a significant collective dose.^4^ For a correct application of the justification and optimization principles of radiation protection, it is therefore important to be able to assess the patient dose for any given CBCT scanner and exposure protocol. While extensive work has been performed to assess organ and effective dose (ED) for CBCT using physical measurements^5–8^ or Monte Carlo simulations^9–11^ on anthropomorphic phantoms, the most convenient and flexible way to estimate dose would be by converting a dose index.

Due to the wide-beam geometry and the highly variable beam collimation in dental CBCT, traditional metrics such as the computed tomography dose index were found to be unsuitable.^12,13^ Whereas modified phantom-based indices have been proposed,^12^ dose-area product (DAP) has become the most commonly used dose index in dental CBCT.^14^ Several countries have established diagnostic reference levels based on DAP,^14^ and it is commonly included in QC protocols as well as for dose monitoring and optimization studies.^15–17^

The main limitation to DAP is its complex relation to patient dose. As DAP is a measurement of tube output rather than the dose absorbed inside a phantom or patient, its conversion to patient dose depends on several factors, primarily the X-ray spectrum and beam geometry. Previous studies estimated conversion coefficients (CC) from DAP to effective dose for dental CBCT;^18,19^ while some of these coefficients were kV and/or FOV-dependent, they may lead to considerable errors in dose estimation for ‘non-average’ CBCT protocols (*e.g.* with a high amount of beam filtration). Another previous study proposed a set of 3 dedicated formulas for the conversion from DAP to effective dose in dental CBCT using multiple linear regression (MLR);^20^ an important improvement was the inclusion of the half-value layer (HVL) to represent beam energy rather than the tube voltage (kV), as well as the separate inclusion of beam width and height. These MLR formulas showed a reduced error for effective dose estimation on the range of exposure parameters found in CBCT compared with the aforementioned CCs. However, due to the relative simplicity of these MLR formulas, it is likely that they do not fully represent the effect of, and interactions between, scan parameters on ED/DAP conversion. The development of more complex conversion models, for example through deep learning involving multi-layer neural networks (NNs), can potentially improve the accuracy of ED/DAP estimation. In addition, for the purpose of dose monitoring, radiobiology research, and epidemiological research, it would be relevant to estimate individual organ doses rather than a direct estimation of the effective dose, as the latter is simply a weighted sum of the former. Thus, the purpose of this study was to develop NN models to estimate organ doses for dental CBCT examinations based on DAP.

## Methods

### Monte Carlo simulation of CBCT dose

CBCT exposures of the dentomaxillofacial region of an adult hermaphrodite phantom (178.6 cm, 73.2 kg) were simulated using PCXMC 2.0 (STUK, Finland).^21^ Based on pilot simulations, CBCT exposures were simulated using 45 projections at 8° intervals, with 20000 simulated photons per projection.^20^ The simulated protocols covered the full range of scan parameters found in clinical CBCT. Two types of protocols were simulated:^20^

- Permutations of fixed scan parameters (n=10944): all combinations of the following sets of parameters were simulated:

`o` Tube voltage: between 70 and 120 kV, at 10-kV intervals. Note that certain CBCT scanners have the option to go below 70 kV, but it is assumed that these are non-clinical protocols.
`o` Filtration: Al filtration thicknesses of 2.5, 5.0, 7.5 and 10 mm were simulated. For Al filtrations up to 5.0 mm, Cu filtration of 0.0, 0.25 or 0.5 mm was added. In combination with the range in tube voltage, this ensured that all types of beam energy distributions found in dental CBCT were included.
`o` Field of view (FOV): 57 FOV sizes and positions. FOV sizes ranged from 4 to 29.8 cm beam width, and 4 to 26.7 cm beam height, at isocenter. For FOVs with a beam height ≤6 cm, both upper and lower jaw positions were included. For FOVs with a beam width ≤6 cm, both anterior and posterior positions were included. Note that all FOVs were positioned in accordance with dental examinations; no FOV positions were used for temporomandibular joint, sinus, ear, etc.
`o` Source-isocenter distance (SID): 35, 45, 55 or 65 cm.
- Permutations of random scan parameters (n=13,440): these simulations used the same range for each scan parameter as above, but with randomly determined values rather than fixed intervals, along with certain constraints for FOV size and position to ensure that the scan protocols are ‘dental’.

In total, 24384 protocols were simulated, with an estimated total simulation time of 1320 h (55 days).

Equivalent organ doses (*H_T_*) for each organ listed in ICRP Publication 103 were recorded for each simulation, along with dose-area product (DAP) values.^3^ The following 13 individual organs were included: bone surface, brain, breast, extrathoracic region, lungs, lymph nodes, muscle, oesophagus, oral mucosa, red bone-marrow, salivary glands, skin, and thyroid. In addition, other organs with a negligible or zero contribution to the effective dose (*i.e.* found well outside the head and neck region) were summed together as ‘minor organs’.

### Neural network training

Two types of fully-connected multilayer perceptron-type NNs were built using Keras (v2.4.0), TensorFlow (v2.3.0) and Scikit-learn (v1.0.2) for Python. Both types of NN used all aforementioned scan parameters as input: tube voltage, filtration (mmAl), additional filtration (mmCu), source-isocenter distance, beam diameter, and beam height. The difference between the two types of NNs is found in how the FOV position is defined:

1. ‘*Coordinate*’ mode, which takes the XYZ-coordinates of the isocenter as input;
2. ‘*AP/JAW*’ mode, which takes categorical codes for the anteroposterior (AP) and craniocaudal (JAW) position as input rather than exact coordinates.

For *AP/JAW* mode, the FOV position was encoded as follows:

- ‘*AP*’ +1 for anterior position, -1 for posterior position, 0 for full arch / not applicable.
- ‘*JAW*’: +1 for upper jaw, -1 for lower jaw, 0 for both jaws / not applicable.

All other input parameters were scaled to a mean of 0 and variance of 1 using Scikit-learn’s StandardScaler tool. The output parameters (*i.e.*, the equivalent dose per DAP) was scaled to a mean of 1 for each organ.

Next, the simulated data was randomly split into training/validation/test data using a 3/1/1 split (*i.e.*, 14600/4868/4868 protocols). A seed was used for the random split, to ensure that the data was split the same for all NNs. The training and validation data were used to design a network architecture and optimal set of hyperparameters for NNs for each of the 14 aforementioned organs, for ‘*Coordinate*’ and ‘*AP/JAW*’ modes, resulting in 28 independent NNs. Rather than using a fixed NN design for each organ and mode, each of the 28 NNs was customized to ensure optimal performance and generalizability. To this end, permutations of the following hyperparameter values were considered:

- Dropout rate (0, 0.25, 0.5)
- Learning rate (0.001, 0.0025, 0.005, 0.0075, 0.01, 0.025, 0.05, 0.075, 0.1)
- Number of hidden layers (1, 2, 3, 4)
- Number of units per hidden layer (10, 25, 50, 100; independent for each layer)
- Activation function for hidden layers (relu, softmax, tanh; note that relu was used as activation function for the output layer)
- Batch size (32, 64, 128, 256, 512, 1024, 2048, 4096)

To efficiently determine the optimal combination out of all permutations within this space, the Hyperband algorithm^22^ was used with a maximum number of epochs of 250, a mean squared error loss function and Adam optimizer. On average, determining the optimal set of hyperparameters for a given NN took 1h5m on an Intel Core i5-8265U CPU. The resulting hyperparameters for each NN are found in Tables 1 and 2 for *Coordinate* and *AP/JAW* mode, respectively. Using these hyperparameters, each NN were trained further until a stopping criterion was reached (i.e. when validation loss did not improve for 100 epochs), and the final NN models were evaluated on the test data.

**Table 1.** Network architecture and hyperparameters for each organ for 9-input networks (‘*Coordinate*’ method)

| Organ | Dropout rate | Learning rate | Layers | Units per layer | Activation function | Batch size |
| --- | --- | --- | --- | --- | --- | --- |
| Bone surface | 0.0 | 0.001 | 2 | 100<br>100 | relu | 32 |
| Brain | 0.0 | 0.01 | 2 | 100<br>50 | softmax | 128 |
| Breast | 0.0 | 0.005 | 4 | 100<br>25<br>25<br>50 | relu | 64 |
| Extrathoracic region | 0.0 | 0.01 | 4 | 100<br>10<br>10<br>50 | relu | 128 |
| Lungs | 0.0 | 0.0025 | 2 | 100<br>25 | relu | 64 |
| Lymph nodes | 0.0 | 0.01 | 4 | 100<br>100<br>25<br>25 | relu | 128 |
| Minor organs | 0.0 | 0.01 | 4 | 100<br>100<br>25<br>25 | relu | 128 |
| Muscle | 0.0 | 0.0025 | 4 | 100<br>100<br>10<br>10 | relu | 32 |
| Oesophagus | 0.0 | 0.01 | 4 | 50<br>100<br>10<br>10 | relu | 256 |
| Oral mucosa | 0.0 | 0.005 | 3 | 50<br>100<br>50 | relu | 128 |
| Red bone-marrow | 0.0 | 0.001 | 1 | 100 | relu | 64 |
| Salivary glands | 0.0 | 0.001 | 3 | 50<br>50<br>50 | tanh | 64 |
| Skin | 0.0 | 0.001 | 2 | 100<br>50 | relu | 32 |
| Thyroid | 0.0 | 0.005 | 3 | 100<br>50 | relu | 64 |

*relu*: rectified linear unit, *softmax*: normalized exponential, *tanh*: hyperbolic tangent
<sup>a</sup>For all layers except output layer, for which relu was used

**Table 2.** Network architecture and hyperparameters for each organ for 8-input networks (‘*AP/JAW*’ method)

| Organ | Dropout rate | Learning rate | Layers | Units per layer | Activation function<sup>a</sup> | Batch size |
| --- | --- | --- | --- | --- | --- | --- |
| Bone surface | 0 | 0.005 | 4 | 100<br>10<br>50<br>10 | relu | 32 |
| Brain | 0 | 0.0025 | 1 | 50 | tanh | 32 |
| Breast | 0 | 0.0075 | 2 | 100<br>25 | relu | 128 |
| Extrathoracic region | 0 | 0.0025 | 1 | 100 | softmax | 128 |
| Lungs | 0 | 0.0075 | 4 | 50<br>100<br>50<br>25 | softmax | 256 |
| Lymph nodes | 0 | 0.0075 | 4 | 50<br>10<br>10<br>50 | relu | 128 |
| Minor organs | 0 | 0.01 | 2 | 50<br>100 | softmax | 32 |
| Muscle | 0 | 0.001 | 2 | 100<br>10 | relu | 32 |
| Oesophagus | 0 | 0.0025 | 1 | 50 | tanh | 128 |
| Oral mucosa | 0 | 0.001 | 2 | 50<br>10 | tanh | 32 |
| Red bone-marrow | 0 | 0.0025 | 2 | 100<br>10 | relu | 64 |
| Salivary glands | 0 | 0.005 | 1 | 100 | softmax | 32 |
| Skin | 0 | 0.0075 | 3 | 50<br>10<br>10 | relu | 32 |
| Thyroid | 0 | 0.005 | 2 | 50<br>50 | softmax | 256 |

*relu*: rectified linear unit, *softmax*: normalized exponential, *tanh*: hyperbolic tangent
<sup>a</sup>For all layers except output layer, for which relu was used

### Comparison with existing conversion coefficients/formulas

The performance of the resulting NN model on the test data was compared with that of the aforementioned MLRs model,^20^ as well as that of previously published CCs.^18,19^ To this end, the ED for the NN models was calculated for each of the 4868 test scan protocols as a weighted sum of each NN output using ICRP 103 tissue weighting factors.^3^

The MLR model, which was fitted on the same simulated data (albeit with a different training/validation split), incorporates the HVL (mmAl) as well as the width (*W*, cm) and height (*H*, cm) of the FOV at isocenter. Three formulas were defined^20^ for the following FOV categories:

- Small FOV (<100 cm^2^ beam area at isocenter):

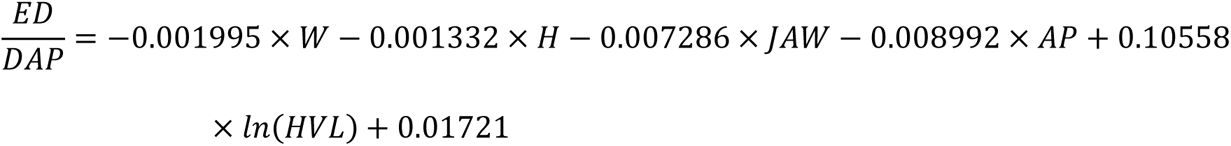

For this formula, similar to the first NN approach used in the current study, the anteroposterior (AP) and craniocaudal (JAW) position is taken into account, using the same numbering system described above.

- Medium FOV (100-400 cm^2^ beam area at isocenter):

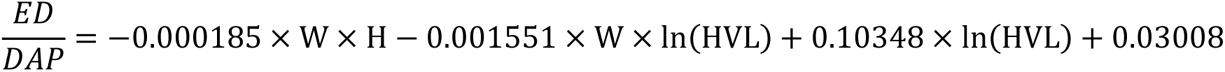

- Large FOV (>400 cm^2^ beam area at isocenter):

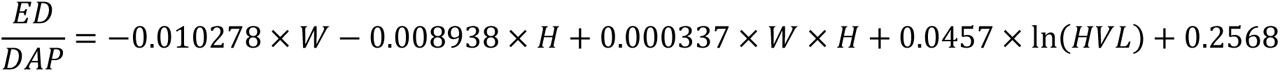

Two sets of CCs were used for comparison with the NNs. First, a kV-dependent coefficient:^19^

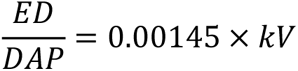

Second, a kV- and FOV-dependent coefficient:^18^

- Small FOV (<100 cm^2^ beam area at isocenter):

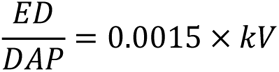

- Medium FOV (100-225 cm^2^ beam area at isocenter):

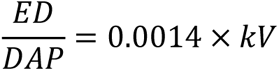

- Large FOV: >225 cm^2^ beam area at isocenter):

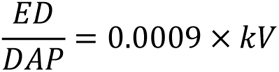

The mean absolute error (MAE, %) between the actual and estimated ED/DAP was calculated for the test data for the two NN-based approaches proposed in this study as well as the MLR model and CCs. Note that the comparison is performed for the effective dose only, as the MLR model or CCs do not yield organ doses.

### Analysis of relative feature importance

To enhance explainability of the NN models, the SHAP (SHapley Additive exPlanations) unified method was used to assess the relative contribution of each input feature to the organ dose from each NN model. The median absolute SHAP values were calculated; in brief, the SHAP values are calculated by assessing the effect of an input feature on a model’s output for a given dataset (in this case, the test data), relative to a given baseline value (i.e. the average output of a model).

## Results

### NN performance

Table 3 shows the range in H_T_/DAP for each organ, as well as ED/DAP, and the MAE for the two NN methods on the test data. The MAE ranged from 0.18% (bone surface) to 2.90% (oesophagus) for the ‘*Coordinate*’ NNs, and from 2.74% (red bone-marrow) to 14.13% (brain) for the ‘*AP/JAW*’ NNs. For 10 out of 14 organs, the MAE for the ‘*Coordinate*’ method was below 1%. For the ‘*AP/JAW*’ method, the MAE was below 10% for 10 out of 14 organs. The MAE for effective dose was 0.23% for ‘*Coordinate*’ mode and 4.32% for ‘*AP/JAW*’ mode.

**Table 3.**
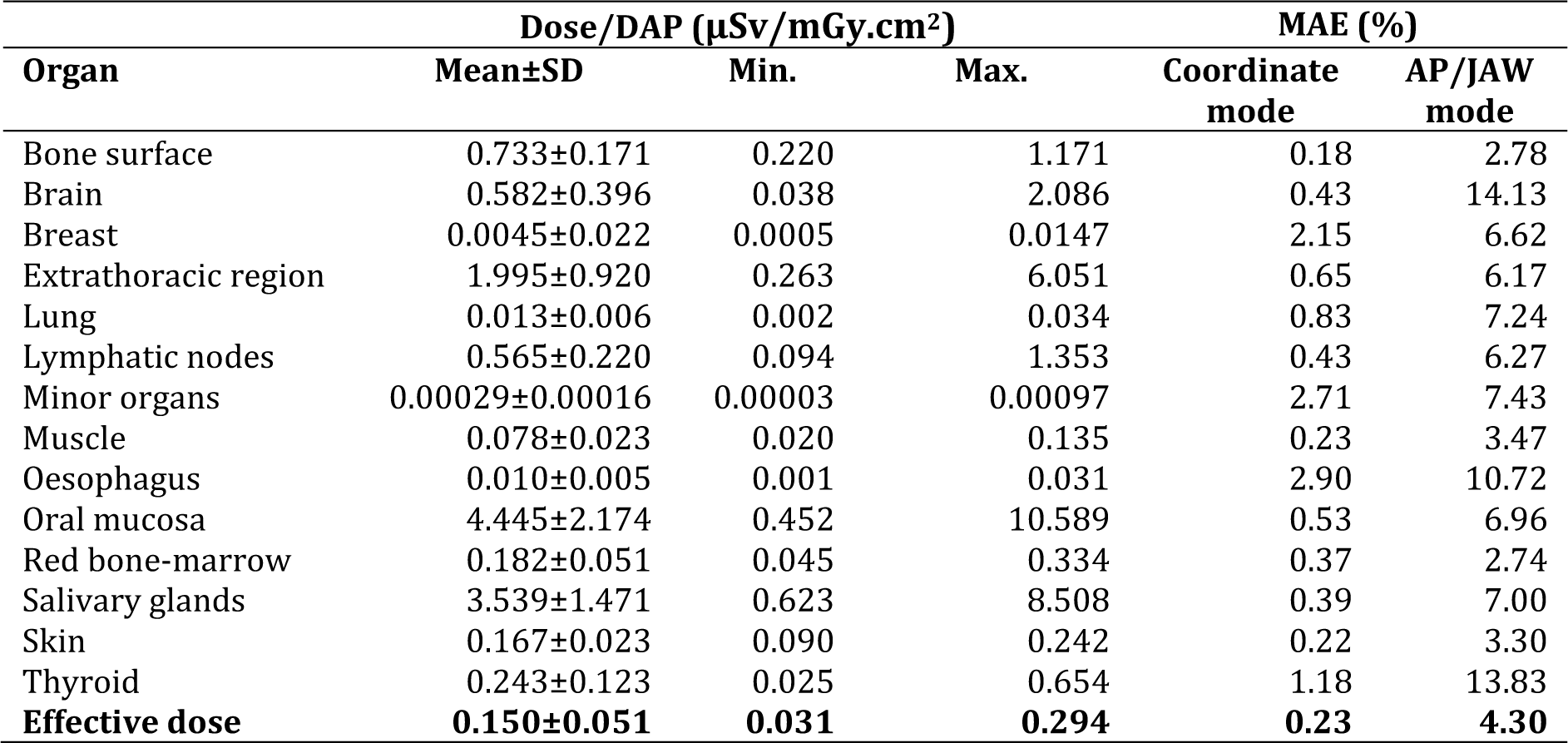
Equivalent/effective dose per dose-area-product (DAP), and mean absolute error (MAE) for the two types of neural networks.

| Organ | Dose/DAP ($\mu Sv/mGy.cm^2$) | | | MAE (%) | |
| --- | --- | --- | --- | --- | --- |
| | Mean±SD | Min. | Max. | Coordinate mode | AP/JAW mode |
| Bone surface | 0.733±0.171 | 0.220 | 1.171 | 0.18 | 2.78 |
| Brain | 0.582±0.396 | 0.038 | 2.086 | 0.43 | 14.13 |
| Breast | 0.0045±0.022 | 0.0005 | 0.0147 | 2.15 | 6.62 |
| Extrathoracic region | 1.995±0.920 | 0.263 | 6.051 | 0.65 | 6.17 |
| Lung | 0.013±0.006 | 0.002 | 0.034 | 0.83 | 7.24 |
| Lymphatic nodes | 0.565±0.220 | 0.094 | 1.353 | 0.43 | 6.27 |
| Minor organs | 0.00029±0.00016 | 0.00003 | 0.00097 | 2.71 | 7.43 |
| Muscle | 0.078±0.023 | 0.020 | 0.135 | 0.23 | 3.47 |
| Oesophagus | 0.010±0.005 | 0.001 | 0.031 | 2.90 | 10.72 |
| Oral mucosa | 4.445±2.174 | 0.452 | 10.589 | 0.53 | 6.96 |
| Red bone-marrow | 0.182±0.051 | 0.045 | 0.334 | 0.37 | 2.74 |
| Salivary glands | 3.539±1.471 | 0.623 | 8.508 | 0.39 | 7.00 |
| Skin | 0.167±0.023 | 0.090 | 0.242 | 0.22 | 3.30 |
| Thyroid | 0.243±0.123 | 0.025 | 0.654 | 1.18 | 13.83 |
| **Effective dose** | **0.150±0.051** | **0.031** | **0.294** | **0.23** | **4.30** |

Scatter plots comparing the true vs. NN-estimated dose are found in Figures 1-6. Each plot depicts the coefficient of determination (R^2^) for a linear fit with an intercept at 0. For ‘*Coordinate*’ mode in particular, a near-perfect correspondence can be seen for most NN models, with R^2^ values between 0.99629 (oesophagus) and 0.99991 (brain) for individual organs and an R^2^ of 0.99991 for effective dose. For ‘*AP/JAW*’ models, minor deviations can be found for most organs (R^2^: 0.92588-0.97887), and somewhat larger deviations for skin (R^2^: 0.85151), thyroid (R^2^: 0.88513) and salivary glands (R^2^: 0.90080), with the effective dose showing an R^2^ of 0.95629.

**Fig. 1.**
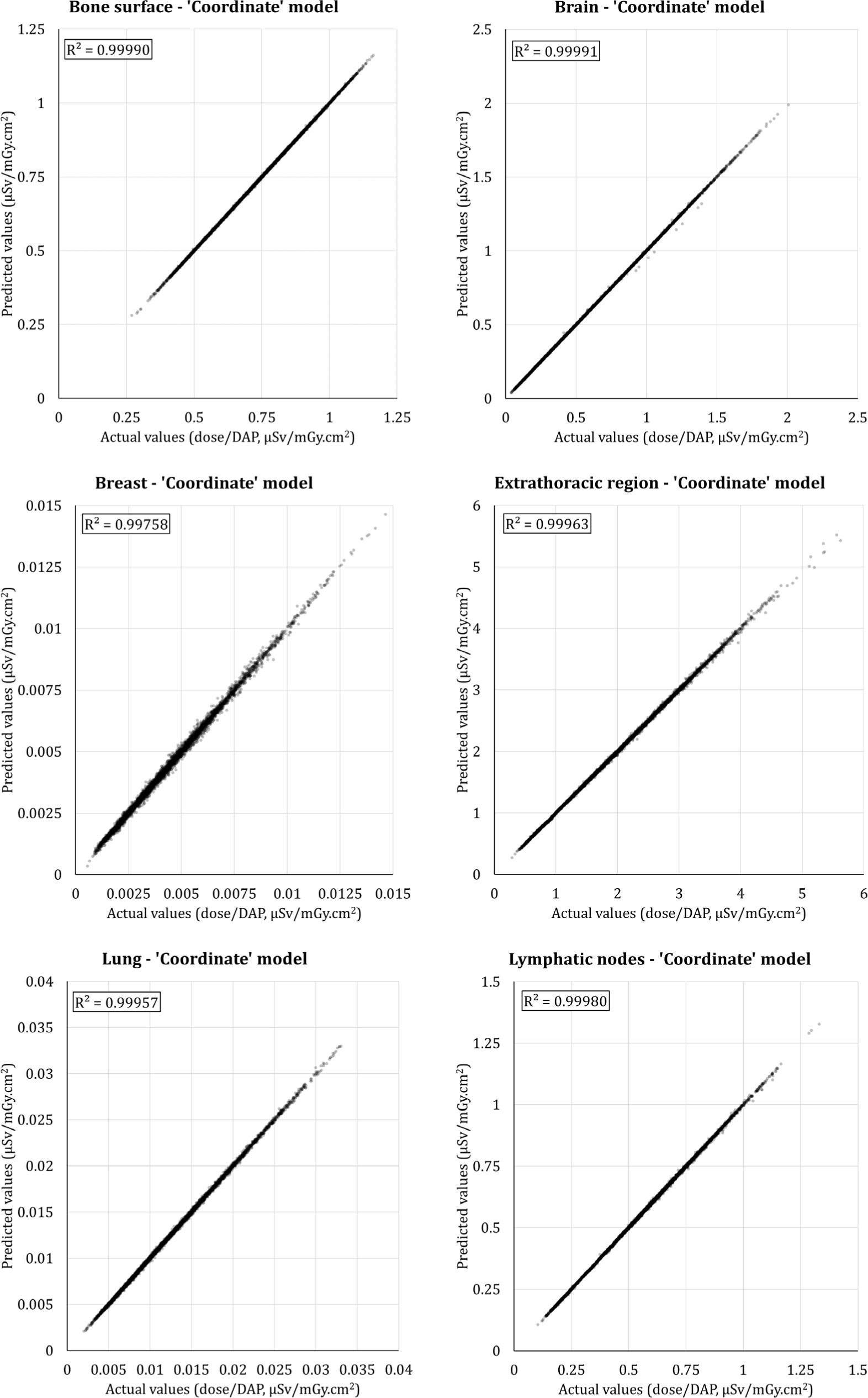
Correlation between actual and estimated equivalent dose per dose-area product (µSv/mGy.cm^2^) for bone surface, brain, breast, extrathoracic region, lung, and lymphatic nodes. Doses are derived from a test dataset comprising 4868 scan protocols and neural network models in ‘*Coordinate*’ mode.

**Fig. 2.**
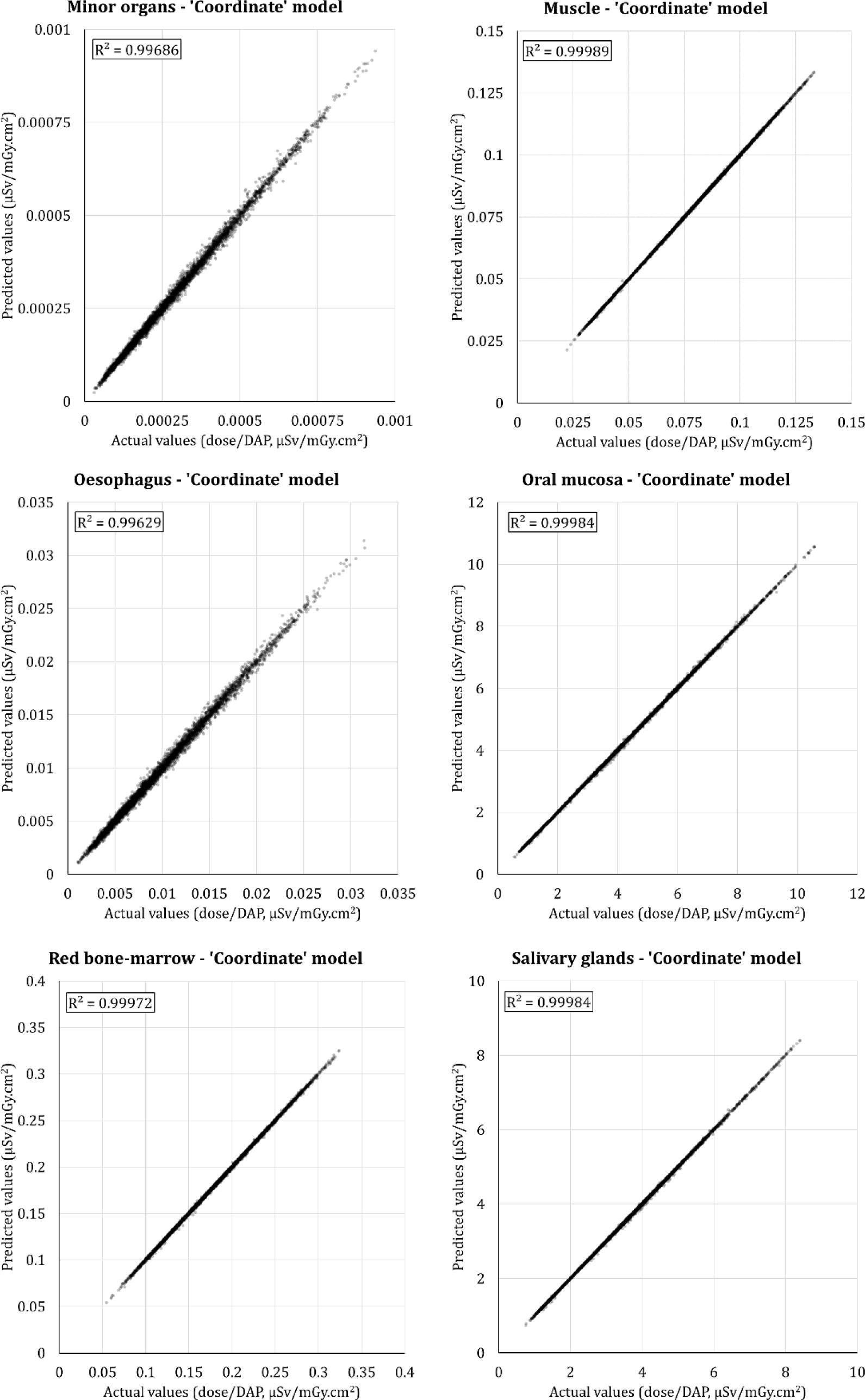
Correlation between actual and estimated equivalent dose per dose-area product (µSv/mGy.cm^2^) for minor organs, muscle, oesophagus, oral mucosa, red bone-marrow, and salivary glands. Doses are derived from a test dataset comprising 4868 scan protocols and neural network models in ‘*Coordinate*’ mode.

**Fig. 3.**
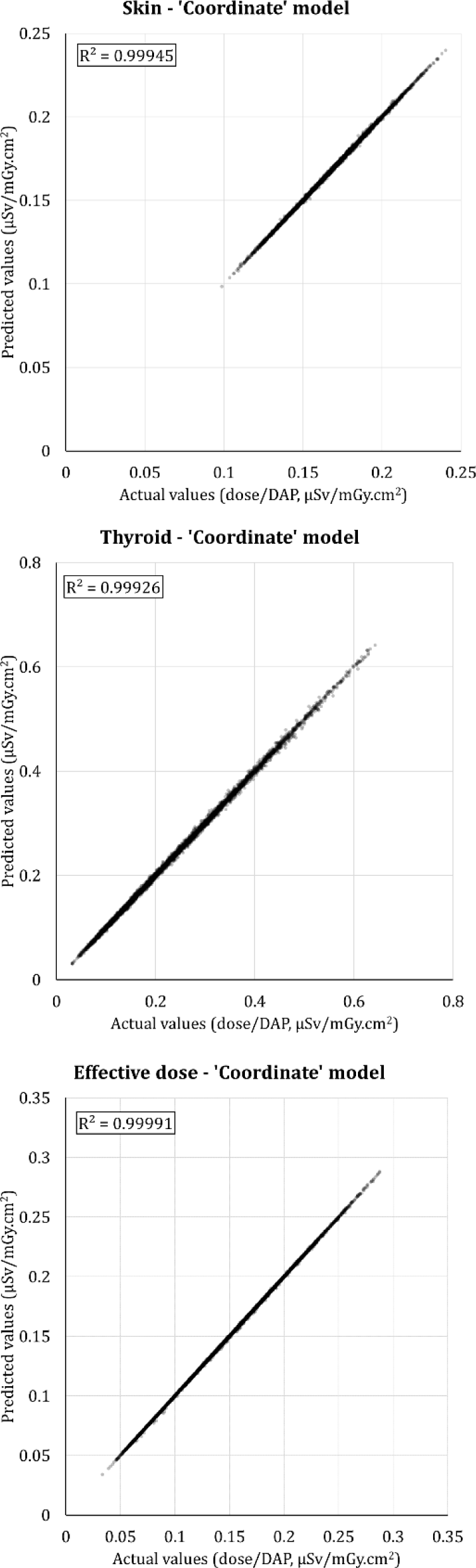
Correlation between actual and estimated equivalent dose (for skin and thyroid) and effective dose per dose-area product (µSv/mGy.cm^2^). Doses are derived from a test dataset comprising 4868 scan protocols and neural network models in ‘*Coordinate*’ mode.

**Fig. 4.**
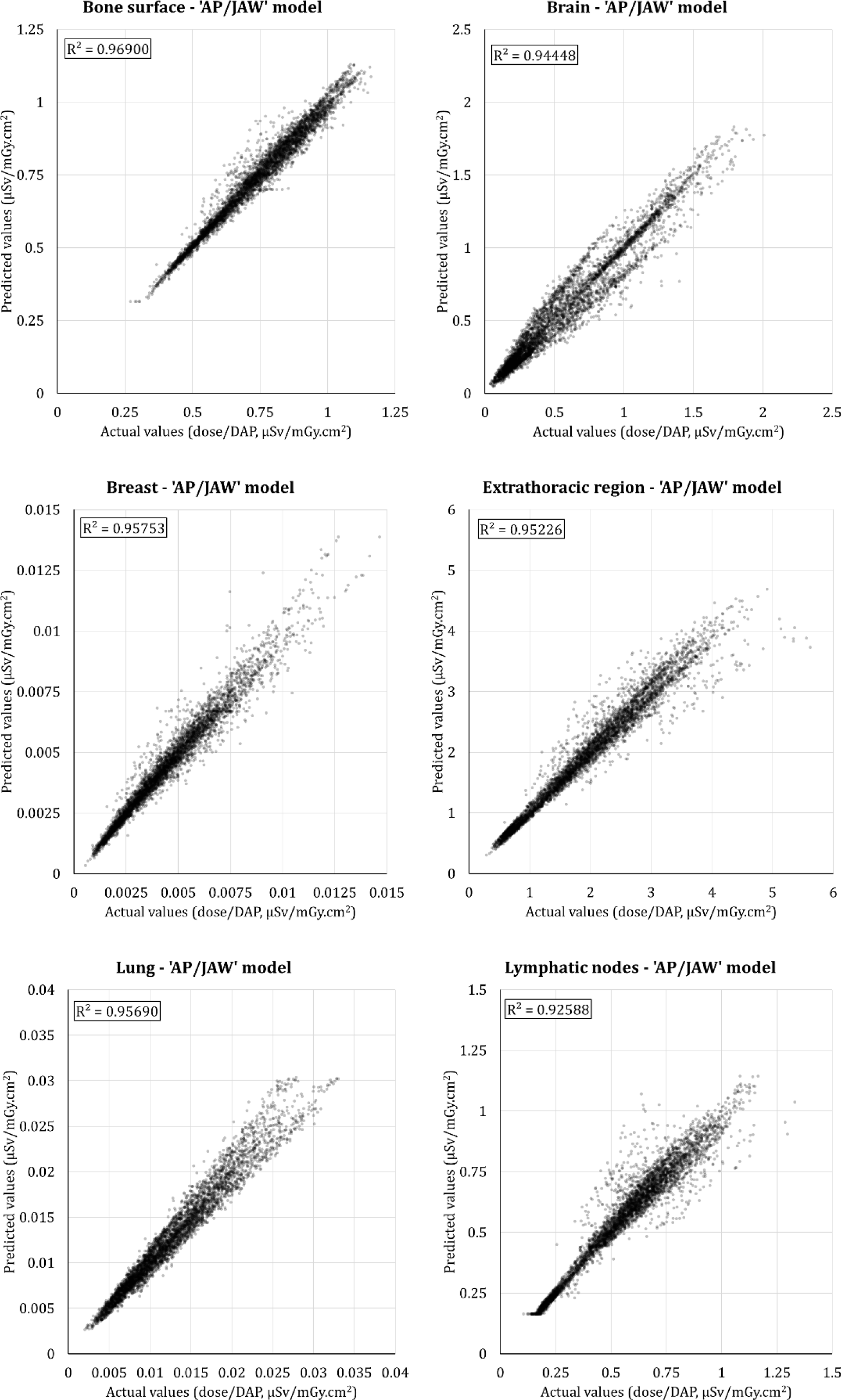
Correlation between actual and estimated equivalent dose per dose-area product (µSv/mGy.cm^2^) for bone surface, brain, breast, extrathoracic region, lung, and lymphatic nodes. Doses are derived from a test dataset comprising 4868 scan protocols and neural network models in ‘*AP/JAW*’ mode.

**Fig. 5.**
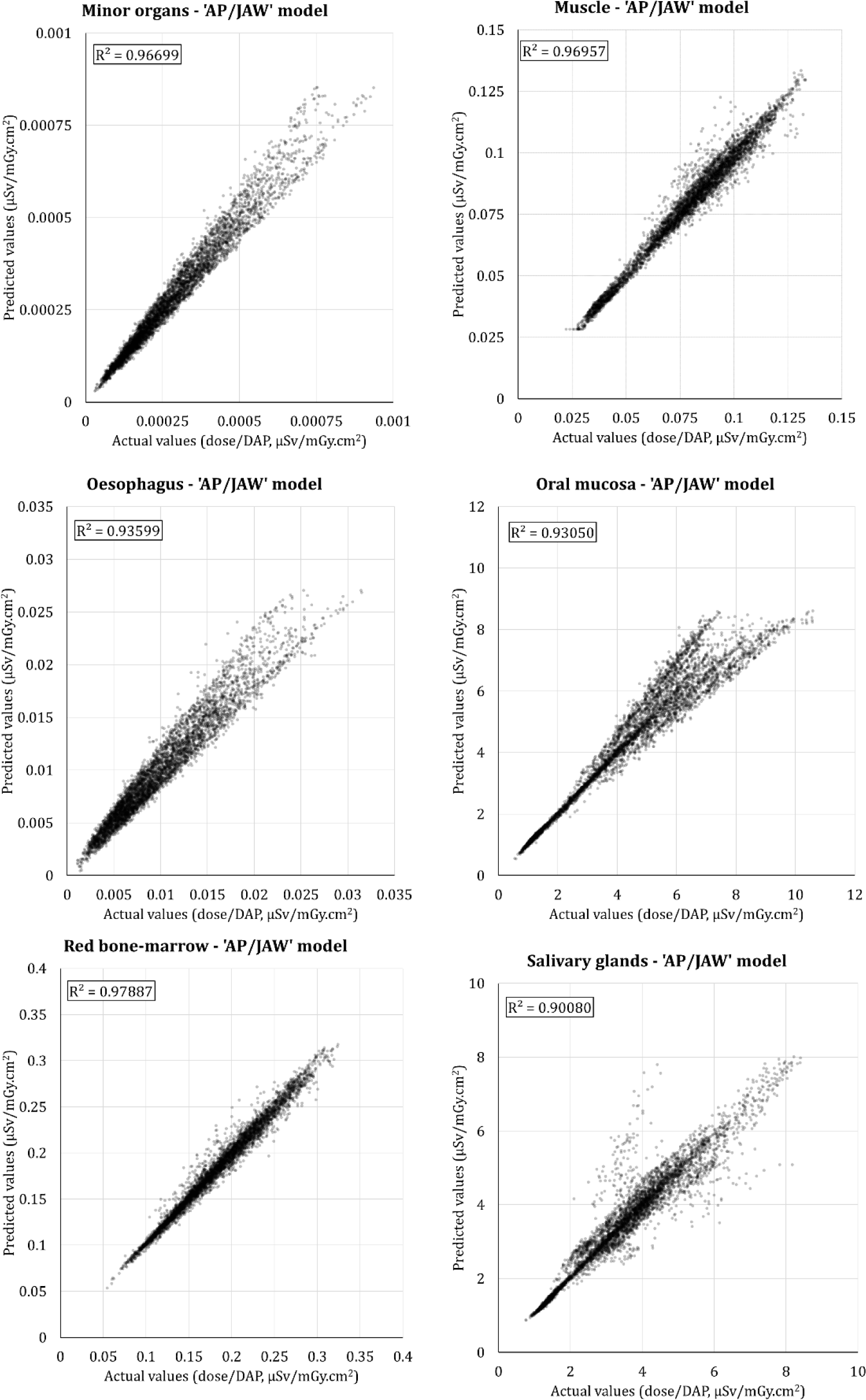
Correlation between actual and estimated equivalent dose per dose-area product (µSv/mGy.cm^2^) for minor organs, muscle, oesophagus, oral mucosa, red bone-marrow, and salivary glands. Doses are derived from a test dataset comprising 4868 scan protocols and neural network models in ‘*AP/JAW*’ mode.

**Fig. 6.**
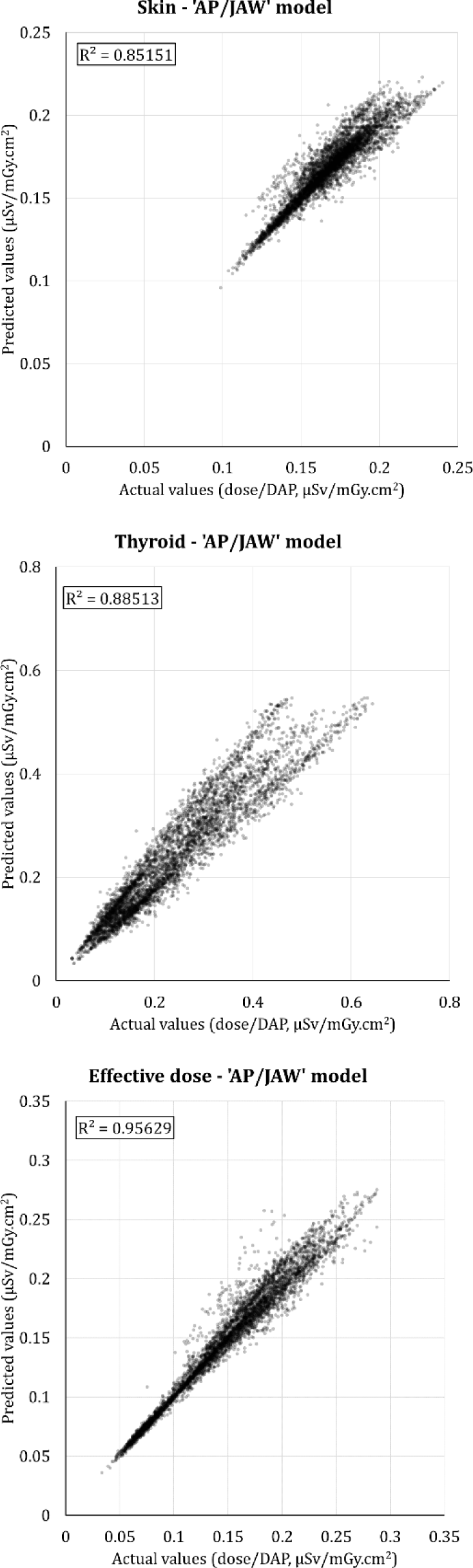
Correlation between actual and estimated equivalent dose (for skin and thyroid) and effective dose per dose-area product (µSv/mGy.cm^2^). Doses are derived from a test dataset comprising 4868 scan protocols and neural network models in ‘*AP/JAW*’ mode.

### Comparison with existing conversion coefficients/formulas

The MAE for ED on the test data was 5.70% for the MLR formulas,^20^ 32.67% for the kV-dependent CC,^19^ and 20.19% for the FOV- and kV-dependent conversion coefficients.^18^

### Analysis of relative feature importance (SHAP)

Figure 7 shows median SHAP values for each input feature and each NN, calculated for the test dataset. Note that feature importance is expressed through absolute values, i.e. no distinction is made between a positive or negative contribution to the dose/DAP. SHAP values should primarily be compared row-wise (i.e., for a given organ). A few observations can be made:

- For most NNs in ‘*Coordinate*’, geometric features (beam dimensions and FOV position) showed a greater relative importance than energy features (kV and filtration). In ‘*AP/JAW*’ mode, this was true for beam dimensions and ‘JAW’ position, whereas ‘AP’ position was of lesser importance.
- In ‘*Coordinate*’ mode, there was no consistency in the relative importance of beam width vs. beam height. For most NNS in ‘*AP/JAW*’ mode, however, beam height was the most important feature among the two.
- In ‘*Coordinate*’ mode, among the three FOV coordinates, the Z-coordinate (indicating the craniocaudal position) showed the highest importance for most NNs. This was especially the case for brain, lungs, oesophagus and thyroid. Between the X- and Y-coordinates, the latter (i.e., the anteroposterior position) was more important than the former (i.e., the left-right position).
- The SID was of minor importance for most NNs.
- Interestingly, for most NNs, Cu filtration was somewhat more important than kV or Al filtration in determining the dose.

**Fig. 7.**
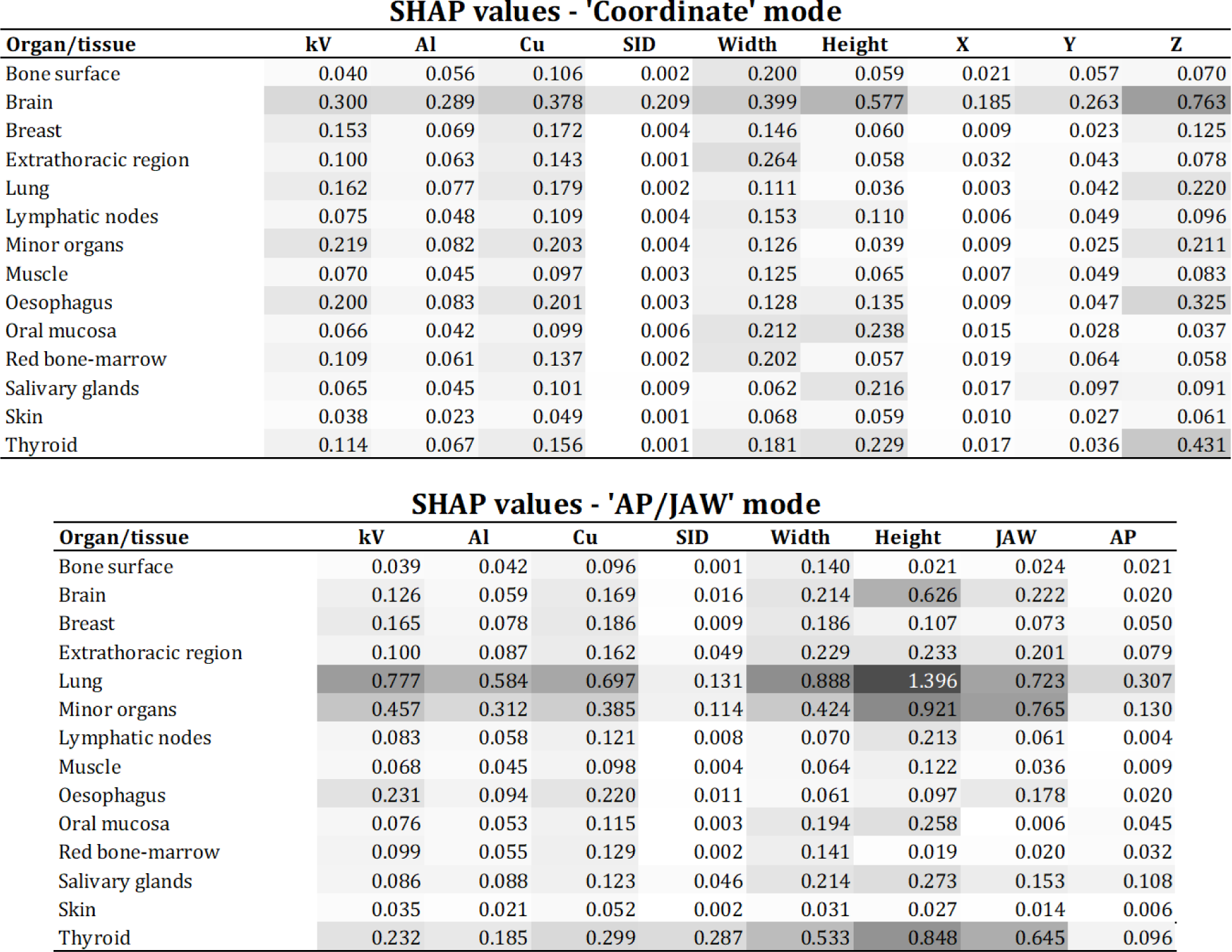
Relative feature importance for neural network models using SHapley Additive exPlanations (SHAP), for each organ/issue and both modes. SHAP values should be compared row-wise (*i.e.* for a given organ), rather than between organs.

## Discussion

In this study, the deep learning was applied to a large dataset of simulated CBCT doses to estimate organ (directly) and effective dose (indirectly) per DAP for a reference patient. Whereas several studies have reported physical or simulated measurements of patient dose in dental CBCT,^5–11^ these are always specific to the scanner model and choice of exposure settings; due to the variety of CBCT scanners on the market,^24^ a standardized dosimetric assessment method based on a dose index is sorely needed. The NN models presented in this study are universally applicable to any CBCT scanner and scan protocol, with a few caveats as described below. Importantly, they allow for the continued use of DAP as a dose index for CBCT.

To the author’s knowledge, this is the first study applying deep learning for the purpose of dental CBCT dosimetry. Deep learning research in medical imaging has focused on several types of tasks, including lesion detection,^25^ segmentation,^26^ and reconstruction.^27^ In radiation therapy, deep learning has been leveraged to provide more accurate treatment plans, e.g. through segmentation or dose prediction.^28^ On the other hand, the use of NNs in patient dosimetry in diagnostic radiology has been scarce. Maier et al. developed a patient-specific dose estimation model using deep learning for CT scanning of the pelvis, abdomen, thorax and head, showing a high correspondence with Monte Carlo simulations at a fraction of the computational time.^29^ Unfortunately, Monte Carlo simulations are far more difficult to set up for dental CBCT, *e.g.* due to FOV truncation and a lack of grey value stability, which complicates the estimation of attenuation coefficients for patient-specific voxel models.

Although both types of NN models (‘*Coordinate*’ and ‘*AP/JAW’*) show an improved dose estimation compared with a conversion formula based on multiple linear regression or a kV/FOV-dependent conversion coefficient, a notable difference in performance could be seen between the two NN types themselves. Whereas most input features are the same for both model types, the use of XYZ coordinates to denote FOV position allowed for a near-perfect dose estimation, whereas the use of categorical positional features resulted in an error of over an order of magnitude higher. This is explained by the fact that exact XYZ-coordinates of the FOV carry a considerably larger amount of information regarding the dose distribution throughout the head and neck than a denotation of the FOV position in terms of the overall dentomaxillofacial region. Previous research has shown that even minor shifts in FOV position can affect organ doses considerably, due to (1) the sharp dose gradients for small FOVs in particular and (2) the presence of radiosensitive organs at the edge of the FOV.^13^ For example, for a FOV categorized as ‘lower jaw’, the dose to the thyroid can vary significantly depending on where the lower edge of the X-ray beam is throughout the scan.

While the ‘*Coordinate*’ models showed superior performance, and could thus be advocated as the models of choice, their practical implementation is more complicated. Whereas both types of models use input features that can be determined in a straightforward manner, the (relative) XYZ-coordinates of a FOV is not readily available for a given scan. One could use indicative coordinates corresponding to typical dental examinations, but this would likely reduce the accuracy of the models to a similar level as the ‘AP/JAW’ ones. In other words, an accurate and patient-specific determination of the XYZ position of the FOV is considered necessary for optimal dose estimation. Although the DICOM Tag ‘(0020,0032) - Image Position (Patient)’ denotes the XYZ-coordinates of the upper left corner of an image,^30^ the author is in doubt whether these values are consistently and accurately reported by CBCT manufacturers, especially because the coordinate frame is also determined by ‘(0010,2210) - Anatomical Orientation Type’. Furthermore, some investigation is needed on how these coordinates are determined; while they are undoubtedly linked to the position of the X-ray tube/detector assembly, it is unclear as of yet how CBCT scanners adapt these values to their positioning tools such as chin/head rests in order to reflect the true position of the patient.

Another way to implement the coordinate-based NNs would be to derive the FOV coordinates from the scan itself, based on the depiction of the patient’s anatomy. Contemporary deep learning-based computer vision approaches could be able to detect certain anatomical landmarks (either on a scout image or a reconstructed scan) and convert these to the reference XYZ coordinates used by the NN models in this study. One can even take it one step further, and use this approach to convert the nominal FOV (based on the actual beam dimensions) to an ‘effective’ FOV (based on the relative size and anatomy of the patient), which would allow the dose to be determined in a patient-specific manner rather than for an adult reference patient. Such an approach will be explored in future work.

A distinct advantage of the NN approaches presented in this study is that they yield equivalent doses for each organ rather than a singular effective dose value. This is particularly relevant for dental exposures, as the dose to each organ (and its relative contribution to the effective dose) can vary considerably, mainly as an effect of FOV size and position. For the data used to develop the NN models in this study, a high variance in relative contribution (%) to the effective dose was seen for the oral mucosa (25.8%±6.8%), brain (5.1%±4.9%) and salivary glands (23.4%±4.7%) in particular.^20^ The ability to estimate equivalent organ doses is pivotal as it allows for a more specific risk estimation; it will be of particular use in radiobiological and epidemiological research involving this modality.

There are a few specific limitations to the dosimetric data on which the NNs were fitted and tested. First, the simulations were performed on an adult reference phantom; extension of the NNs to pediatric exposures would require a similarly sized dataset of simulated CBCT scans on phantoms of varying size, followed by the training of new NNs that take features such as age and gender into account. Alternatively, the concept of an ‘effective FOV size’ mentioned above may be sufficient to adapt the NN for patients of any size with reasonable accuracy. Regarding the accuracy of the simulated doses, whereas the self-assessed uncertainty of the effective dose by PCXMC was around ∼1% for the simulations in this study, it should be noted that the PCXMC phantom model is lacking some detail compared to e.g. the ICRP mesh-type phantoms (although the latter are rudimentary in terms of dental anatomy as well).^31^ Regardless, a high uncertainty should always be considered when applying doses from reference phantoms to individual patients.

Another limitation to the dosimetric set-up is that all dose simulations involved 360° rotations. However, since the NNs’ output is defined as the dose per DAP, they are applicable to 180° rotation or other partial rotation arcs as well. This is supported by previous studies that have shown a limited effect of the rotation angle on the dose per mAs, indicating that the dose is primarily dictated by the total tube output.^32,33^ Regardless, a more elaborate NN model that takes into account the rotation arc (and starting angle / rotation direction) will be considered in future work. A final limitation is that all simulations involved a flat filtration, a symmetric beam geometry and an anode angle of 16°. Uncertainties and possible corrections of the dose estimates for non-flat filtrations, off-axis X-ray beams, and different anode angles remain to be determined.

In conclusion, the NNs developed in this study allow for the accurate estimation of organ and effective dose of CBCT scans for any combination of scan parameters, considering an adult reference patient. Further study should focus on expanding this approach to allow for individual (i.e. size-specific) dosimetry, including for pediatric patients.

## Data Availability

All data produced in the present study are available upon reasonable request to the authors.

## Acknowledgments

R. Pauwels was supported by the European Union Horizon 2020 Research and Innovation Program under the Marie Skłodowska-Curie Grant agreement number 754513 and by Aarhus University Research Foundation (AIAS-COFUND).

## References

1. Pauwels R. Cone beam CT for dental and maxillofacial imaging: dose matters. Radiat Prot Dosimetry. 2015; 165:156–161.

2. Benavides E, Krecioch JR, Connolly RT, Allareddy T, Buchanan A, Spelic D, et al. Optimizing radiation safety in dentistry: Clinical recommendations and regulatory considerations. J Am Dent Assoc. 2024; 155:280–293.e4.

3. The 2007 Recommendations of the International Commission on Radiological Protection. ICRP publication 103. Ann ICRP. 2007; 37:1–332.

4. Benn DK, Vig PS. Estimation of x-ray radiation related cancers in US dental offices: Is it worth the risk? Oral Surg Oral Med Oral Pathol Oral Radiol. 2021; 132:597–608.

5. Pauwels R, Beinsberger J, Collaert B, Theodorakou C, Rogers J, Walker A, et al. Effective dose range for dental cone beam computed tomography scanners. Eur J Radiol. 2012;81:267–271.

6. Vogiatzi T, Menz R, Verna C, Bornstein MM, Dagassan-Berndt D. Effect of field of view (FOV) positioning and shielding on radiation dose in paediatric CBCT. Dentomaxillofac Radiol. 2022; 51:20210316.

7. Rottke D, Patzelt S, Poxleitner P, Schulze D. Effective dose span of ten different cone beam CT devices. Dentomaxillofac Radiol. 2013; 42:20120417.

8. Ludlow JB. A manufacturer’s role in reducing the dose of cone beam computed tomography examinations: effect of beam filtration. Dentomaxillofac Radiol. 2011; 40:115–122.

9. Zhang G, Pauwels R, Marshall N, Shaheen E, Nuyts J, Jacobs R, et al. Development and validation of a hybrid simulation technique for cone beam CT: application to an oral imaging system. Phys Med Biol. 2011; 56:5823–5843.

10. Ozaki Y, Watanabe H, Kurabayashi T. Effective dose estimation in cone-beam computed tomography for dental use by Monte-Carlo simulation optimizing calculation numbers using a step-and-shoot method. Dentomaxillofac Radiol. 2021; 50:20210084.

11. Kim EK, Han WJ, Choi JW, Battulga B. Estimation of the effective dose of dental cone-beam computed tomography using personal computer-based Monte Carlo software. Imaging Sci Dent. 2018; 48:21–30.

12. ICRP; Rehani MM, Gupta R, Bartling S, Sharp GC, Pauwels R, et al. Radiological Protection in Cone Beam Computed Tomography (CBCT). ICRP Publication 129. Ann ICRP. 2015; 44:9–127.

13. Pauwels R, Theodorakou C, Walker A, Bosmans H, Jacobs R, Horner K, et al. Dose distribution for dental cone beam CT and its implication for defining a dose index. Dentomaxillofac Radiol. 2012; 41:583–593.

14. International Atomic Energy Agency, Radiation Protection in Dental Radiology, Safety Reports Series No. 108, IAEA, Vienna, 2022. Available from: https://www.iaea.org/publications/14720/radiation-protection-in-dental-radiology

15. de Las Heras Gala H, Torresin A, Dasu A, Rampado O, Delis H, Hernández Girón I, et al. Quality control in cone-beam computed tomography (CBCT) EFOMP-ESTRO-IAEA protocol (summary report). Phys Med. 2017; 39:67–72.

16. Brasil DM, Pauwels R, Coucke W, Haiter-Neto F, Jacobs R. Image quality optimization using a narrow vertical detector dental cone-beam CT. Dentomaxillofac Radiol. 2019; 48:20180357.

17. Brasil DM, Merken K, Binst J, Bosmans H, Haiter-Neto F, Jacobs R. Monitoring cone-beam CT radiation dose levels in a University Hospital. Dentomaxillofac Radiol. 2023; 52:20220213.

18. Mah E, Ritenour ER, Yao H. A review of dental cone-beam CT dose conversion coefficients. Dentomaxillofac Radiol. 2021; 50:20200225.

19. Batista WO, Navarro MV, Maia AF. Effective doses in panoramic images from conventional and CBCT equipment. Radiat Prot Dosimetry. 2012; 151:67–75.

20. Pauwels R. A new formula for converting dose-area product to effective dose in dental cone-beam computed tomography. Phys Med. 2023;112:102639.

21. Tapiovaara M, Siiskonen T. PCXMC - A Monte Carlo program for calculating patient doses in medical x-ray examinations. 2nd Ed. Finland: Radiation and Nuclear Safety Authority; 2008.

22. Li L, Jamieson K, DeSalvo G, Rostamizadeh A, Talwalkar A. Hyperband: A Novel Bandit-Based Approach to Hyperparameter Optimization. arXiv 2016: 1603.06560.

23. Lundberg S, Lee SI. A Unified Approach to Interpreting Model Predictions. arXiv 2017: 1705.07874.

24. Gaêta-Araujo H, Alzoubi T, Vasconcelos KF, Orhan K, Pauwels R, Casselman JW, et al. Cone beam computed tomography in dentomaxillofacial radiology: a two-decade overview. Dentomaxillofac Radiol. 2020; 49:20200145.

25. Mohammad-Rahimi H, Motamedian SR, Rohban MH, Krois J, Uribe SE, Mahmoudinia E, et al. Deep learning for caries detection: A systematic review. J Dent. 2022; 122:104115.

26. Minaee S, Boykov Y, Porikli F, Plaza A, Kehtarnavaz N, Terzopoulos D. Image Segmentation Using Deep Learning: A Survey. IEEE Trans Pattern Anal Mach Intell. 2022; 44:3523–3542.

27. Minnema J, Ernst A, van Eijnatten M, Pauwels R, Forouzanfar T, Batenburg KJ, et al. A review on the application of deep learning for CT reconstruction, bone segmentation and surgical planning in oral and maxillofacial surgery. Dentomaxillofac Radiol. 2022; 51:20210437.

28. Meyer P, Noblet V, Mazzara C, Lallement A. Survey on deep learning for radiotherapy. Comput Biol Med. 2018; 98:126–146.

29. Maier J, Klein L, Eulig E, Sawall S, Kachelrieß M. Real-time estimation of patient-specific dose distributions for medical CT using the deep dose estimation. Med Phys. 2022; 49:2259–2269.

30. National Electrical Manufacturers Association (NEMA). Digital Imaging and Communications in Medicine (DICOM) Part 3: Information Object Definitions. Rosslyn, VA, USA: NEMA; 2023. Available from: https://www.dicomstandard.org/

31. Kim CH, Yeom YS, Petoussi-Henss N, Zankl M, Bolch WE, Lee C, et al. ICRP Publication 145: Adult Mesh-Type Reference Computational Phantoms. Ann ICRP. 2020; 49:13–201.

32. Pauwels R, Zhang G, Theodorakou C, Walker A, Bosmans H, Jacobs R, et al. Effective radiation dose and eye lens dose in dental cone beam CT: effect of field of view and angle of rotation. Br J Radiol. 2014; 87:20130654.

33. Zhang G, Marshall N, Bogaerts R, Jacobs R, Bosmans H. Monte Carlo modeling for dose assessment in cone beam CT for oral and maxillofacial applications. Med Phys. 2013; 40:072103.

